# Universal Health Coverage and COVID-19 Pandemic: A Bangladesh Perspective

**DOI:** 10.1101/2020.11.11.20229526

**Authors:** Most. Zannatul Ferdous, Ummay Soumayia Islam

**Affiliations:** Department of Public Health and Informatics, Jahangirnagar University, Savar, Dhaka-1342

**Author notes:** **Corresponding author** Most. Zannatul Ferdous, Lecturer, Department of Public Health and Informatics, Jahangirnaagar University, Savar, Dhaka-1342. **Authors Contributions** Most. Zannatul Ferdous: Conceptualization, Writing-original draft, Editing, and Validation; Ummay Soumayia Islam: Writing-original draft, Editing, and Validation.

## Abstract

**Background:** Like many other countries around the world, Bangladesh adopts Universal Health Coverage (UHC) as a national aspiration. The central theme of its providing quality and affordable health services which is a significant element of social protection. This paper was aimed to provide a narrative understanding of the perspectives of UHC in Bangladesh towards COVID-19 based on the existing literature.

**Methods:** We conducted a review combining articles and abstracts with full HTML and PDF format. We searched Google Scholar, ScienceDirect and Google using multiple terms related to UHC, COVID-19 and Bangladesh without any date boundary and without any basis of types of studies, that is, all types of studies were scrutinized.

**Results:** This short description highlights that the current pandemic COVID-19 holds lessons that health systems and economies in several countries like Bangladesh are not in enough preparation to tackle a massive public health crisis. It reports the shortage of health workers, scarcity of personal protective equipment, limited and ineffective diagnostic facilities, inadequate infrastructure of health care facilities, scarcity of drugs, and underfunded health services. Further, COVID-19 pandemic highlights the country’s health system needs an ongoing rehab post-COVID-19 with strong coordination in governance, in health economics, in health systems, in information systems, as well as in community participation in health to achieve UHC.

**Conclusions:** Addressing the needs for UHC achievement, it is important to break down the access barriers and keeping up to date all the activities addressing public health crisis like COVID-19.

## Perspectives

World Health Organization (WHO) defines Universal Health Coverage (UHC) as- ensuring access to needed health care services including health promotion, prevention, treatment, and rehabilitation when and where appropriate without financial hardship among the user (WHO, 2020a).Approximately half of the people across the globe are deprived from health services they need including 100 million are pushed below poverty line every year because of out of pocket payment on their health[1]. In 2015, the nations of the world set UHC as one of the target under goal 3 when adopting Sustainable Development Goals. By achieving UHC countries can achieve progress towards other health-related targets including other goals since sound health allows children to learn, adults to earn, helps to reduce poverty, and strengthen economic development[2].Therefore, all countries has given their consent towards achieving UHC as part of the 2030 Agenda except Bangladesh. According to the commitment of the Prime Minister of Bangladesh in the 64^th^ World Health Assembly held in May 2011, the country committed to gain UHC by 2032[3]. Less than 1% of the total population has a health coverage scheme against health expenditure including 3.8% is pushed into poverty line for paying health services each year. Though Bangladesh has achieved significant progress over the last 2 decades, 64% health expenditure is still come from out of pocket. As a result it is experiencing the highest expenditure (15%) compared to neighboring country India (10% to 12%) and Thailand (2%)[4].

The recent COVID-19 is an ongoing global public health crisis according to WHO declaration on 30^th^ January, 2020 which was first emerged at Wuhan city of China[5], [6]. In Bangladesh, Institute of Epidemiology, Disease Control and Research (IEDCR) declared the first confirmed cases on 8^th^ March, 2020 [7]. After that country has taken numerous control measures fighting against COVID-19 though these available control measures are significantly influenced by the knowledge, attitudes, and practices (KAP) towards COVID-19[8]. However, adoption of control measures, the country is facing frequent problem like economic and social standstill and among them health sector is the most affected sector by the pandemic[9]. As of 9^th^ November, 2020 Bangladesh confirmed 42,238 cases with 6067 deaths and 338,145 recovery [10]. This study aims to assess the impact of COVID-19 in Bangladesh and lesson learned towards achieving UHC by 2032.

### Literature search strategies

We reviewed the literature (PubMed, Google Scholar, ScienceDirect, Google and online newspapers in Bangladesh) to get recent information on UHC, UHC and COVID-19 in Bangladesh.

**Figure 1:**
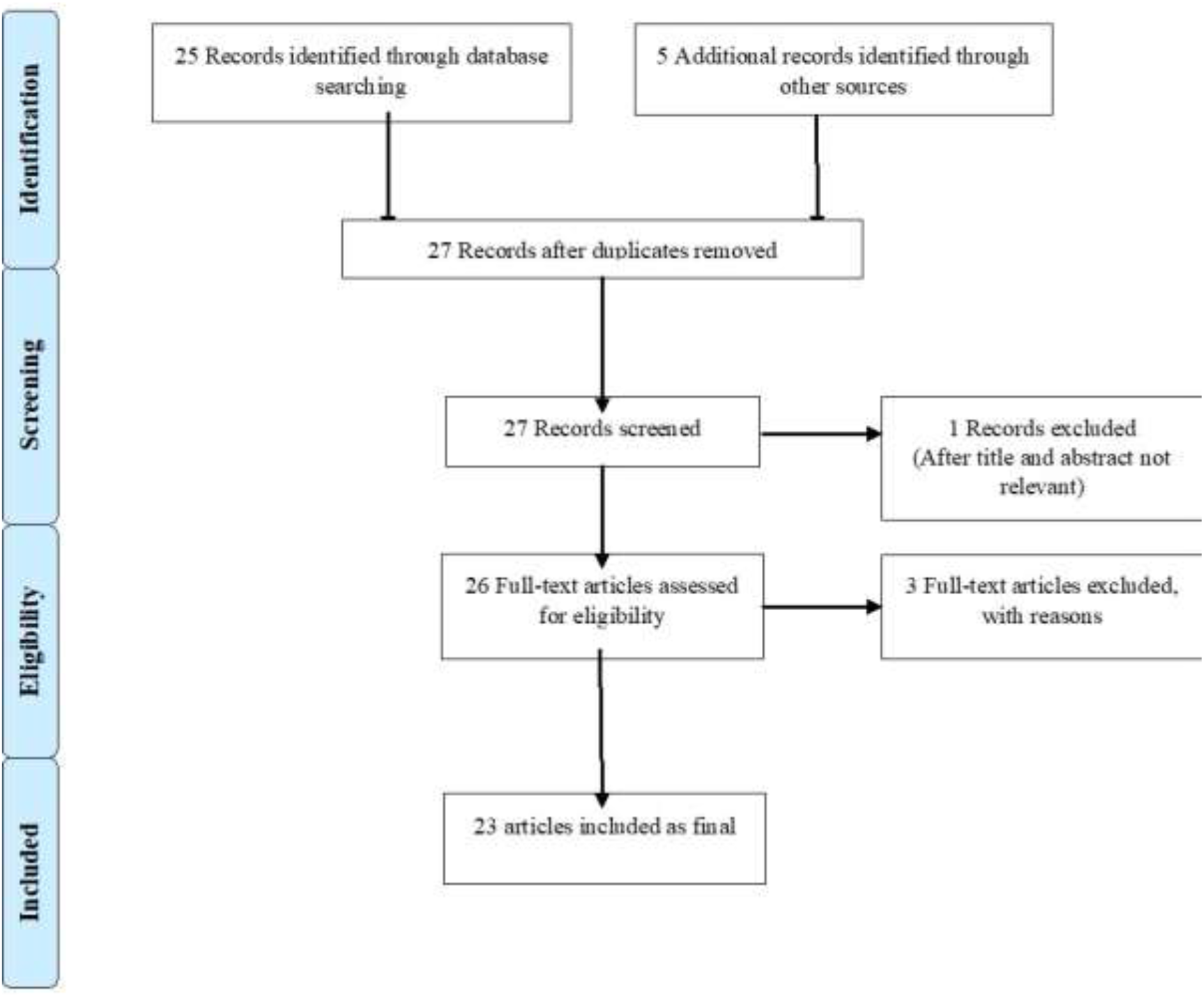
Flow diagram of the study selection process (PRISMA flow diagram)

### Lessons towards COVID-19 and Bangladesh

Public health emergency leads any country in a danger if it beyond under control. Likewise, recently declared public health emergency COVID-19 affected more than 216 countries and territories with 50,782,732 confirmed cases and 1,262,751 deaths as of 9^th^ November, 2020 around the world. It has shown the critical need for the preparedness during a disaster to the world. Preparation is the fundamental tool for mitigating the impact of any catastrophe and COVID-19 taught the world a great lesson by showing its severity. When many countries are struggling to provide UHC to its community, a new emerging public health problem such as COVID-19 pose a global threat and continue to rise and spread its impact on people in many ways like physically, mentally, economically, and so on. Extended health emergencies due to the pandemic of COVID-19 the supply sides (like health systems) went under an enormous pressure. The aftermath has a significant impact on the vulnerable population by lacking social protection and access to health care services in their normal life [11]. The Lancet Global Health Commission addressed poor quality care is now a bigger barrier to reducing mortality than insufficient access. It estimates approximately 60% of deaths from conditions amenable to health care are due to poor quality care, whereas the remaining deaths result from non-utilization of the health system[12]. This situation more is dreadful for vulnerable groups including the poor, the less educated, adolescents, those with stigmatized conditions, and those at the edges of health systems, such as people in prison [13]. Compared to other South Asian countries, Bangladesh has one of the best government health infrastructure including 500 upazila health complex, 5000 union sub centers, 13000 satellite clinics, secondary, and tertiary care hospitals. Yet the country is facing lack of skilled health care professionals particularly in rural areas, scarcity of health care resources, financial constraints. Most of the time patients have to pay all cost related to health service from their pocket which leads to poor people become poorer and even rich are also facing hardship with the costly treatment procedure[4]. During COVID-19 pandemic the country has a severe scarcity of testing kits in stocks and only some 20,000 have been distributed to other parts testing laboratories around the country[14], [15]. However, China provide some testing kits, masks, infrared thermometers, and PPE to deal with the crisis but this amount only covers a small proportion needs[16].

Approximately, 1.1 million slum dwellers living in extremely close quarters are hardly conscious about the threat of COVID-19 and most of them passed their life with great difficulty by losing jobs due to pandemic crisis. These families have less facilities of bathroom, toilet with scarcity of water. Therefore, maintaining hygiene is a challenge to them. The same situation goes for Rohingya refugees, who fled in Bangladesh from neighboring country Myanmar in 2017, leading to vulnerable health condition among these population and increase more chance to illness[17-19]. Since there is scarcity of PPE (personal protective equipment), testing facilities most of the health workers which is not sufficient compared to large population refused to provide service with this crisis due to fear of infection[20]. According to a study conducted in Gazipur Upazila, e-health care have dropped 80% due to absence of doctors, 58% being deprived of basic human needs, 71% pregnant women missed their regular ANC check-ups, 70% female faced familial complications, and Expanded Program on Immunization (EPI) has also dropped during pandemic [21]. As a result, shortage of health workers, scarcity of personal protective equipment, inadequate infrastructure of health care facilities, scarcity of drugs, and underfunded health services has revealed more clearly during this pandemic that shows a consistent impediment to achieve UHC in Bangladesh. In addition relocating medical staff to emergency site from previous department increase scarcity of service delivery in those site like non-communicable diseases, HIV, Tuberculosis, mental health, maternal and child health, increased unwanted pregnancy, malaria, and diphtheria[9]. Furthermore, in this critical time, smart planning with sufficient preparation for mitigating the incidence and prevalence of disease including designing the future prospect is very important to achieve UHC[22]. Because evidence on the management approaches of current COVID-19 pandemic is still limited though the numbers of affected countries are increasing as the days go by[23].

However, to achieve UHC in Bangladesh should include development of a long term national human resources policy and action plan considering the emerging and re-emerging health risks, establishment of a national insurance system, building of an interoperable electronic health information system and investment to strengthen the capacity of the ministry of health.

## Conclusion

Health is a fundamental human right, and UHC is a key to achieve health for all. Addressing the needs for UHC achievement, it is important to break down the access barriers by removing financial, geographical and cultural barriers in a sustained way that reduce the out of pocket expenditure among Bangladeshi people. Additionally, COVID-19 pandemic focused the necessity of healthy economy by controlling the pandemic effectively with sustainable solution measures. As a lower middle income country, Bangladesh has limited resources in health sectors which needed rapid reforms of resources for achieving UHC by 2032. Hence, by developing the need basis stronger and resilient health infrastructure the country will be able to provide timely response during any health crisis including better protection against future outbreak control with access to needed health care excluding financial burden.

## Data Availability

Data are not available to authors. Authors used secondary data sources.

## Competing interests

The authors declare no competing interests.

## Notes

### Competing Interest Statement

The authors have declared no competing interest.

### Funding Statement

Self funded.

### Author Declarations

Data obtained from secondary source, no formal ethical assessment was required.

